# Mental health in the UK during the COVID-19 pandemic: early observations

**DOI:** 10.1101/2020.05.14.20102012

**Authors:** Ru Jia, Kieran Ayling, Trudie Chalder, Adam Massey, Elizabeth Broadbent, Carol Coupland, Kavita Vedhara

**Author notes:** Author to whom all correspondence should be addressed: Professor Kavita Vedhara, Division of Primary Care, University of Nottingham, University Park, Nottingham, NG7 2RD, UK.

## Abstract

**Background:** Previous pandemics have resulted in significant consequences for mental health. Here we report the mental health sequela of the COVID-19 pandemic on the UK population and examine modifiable and non-modifiable explanatory factors associated with mental health outcomes. We focus on the short-term consequences for mental health, as reported during the first four-six weeks of social distancing measures being introduced.

**Methods:** A community cohort study was conducted with adults aged≥18 years recruited through a mainstream and social media campaign between 3/4/20-30/4/20. Consenting participants completed an online survey measuring depression, anxiety and stress and explanatory variables hypothesised to be related to these mental health outcomes.

**Outcomes:** N = 3097 eligible individuals participated. The cohort was predominantly female (85%); mean age forty-four years; 10% from minority ethnic groups; 50% described themselves as key-workers and 20% identified as having clinical risk factors putting them at increased risk of COVID-19. Mean scores for depression, stress and anxiety significantly exceeded population norms. Analysis of non-modifiable factors indicated that being younger and female were associated with all outcomes, with the final multivariable models accounting for 7–13% of variance. When adding modifiable factors, significant independent effects emerged for positive mood, perceived loneliness and worry about getting COVID-19 for all outcomes, with the final multivariable models accounting for 54–57% of variance.

**Interpretation:** Increased psychological morbidity was evident in this UK cohort, with younger people and women at particular risk. Interventions targeting perceptions of: loneliness, risk of COVID-19, worry about COVID-19, and positive mood may be effective.

## Background

The COVID-19 (Coronavirus, 2019) pandemic has resulted in unprecedented disruption to the fabric of society, our health service and economy. However, the multitude of challenges presented by the pandemic may also pose a significant threat to our psychological health.^1^ Individuals are facing a panoply of stressors including: serious illness, bereavement, social distancing, and unemployment. The consequences of these stressors for mental health will not be uniform, rather they will be influenced by a range of modifiable and non-modifiable factors. Understanding these factors will be critical in determining who is at greatest risk of mental health difficulties and potential approaches to intervention. We report here cross-sectional findings from a community cohort study designed to capture both the mental health sequela of the COVID-19 pandemic, as well as the modifiable and non-modifiable explanatory factors associated with adverse mental health outcomes. Our focus is on the immediate consequences for mental health, as reported during the first 4–6 weeks of social distancing measures being introduced in the UK.

In keeping with its recent emergence, much remains unknown about COVID-19 and its consequences. However, the expectation is that the consequences for mental health will be profound and far reaching.^1^ Evidence on the impact of the pandemic on people living in China attests to this possibility. For example, one study reported the prevalence of depression and anxiety in medical staff to be over 50% and 44% respectively.^2^ Similarly, a study with a general population sample during Level 1 of the epidemic (classified as a ‘significant’ emergency) revealed that greater than 70% of participants reported moderate or high psychological symptoms of obsessive compulsion, interpersonal sensitivity, phobic anxiety, and psychoticism.^3^

The experience of previous pandemics also suggests that significant mental health consequences are probable. During the Middle East Respiratory Syndrome outbreak, for example, untoward psychological effects occurred rapidly: with symptoms of anxiety and anger evident after brief isolation periods of 2 weeks.^4^ Similarly, the Severe Acute Respiratory Syndrome epidemic was associated with a 31% increase in suicide amongst the over 65s in Hong Kong.^5^

Preliminary evidence from the UK suggests that these experiences may be replicated here. For example, two online surveys conducted in the first week after social distancing measures were introduced, highlighted some early concerns.^1^ One survey was conducted with people with lived experience of mental health problems and stakeholders (n=2198), and the other a nationally representative sample of the general population (n=1099). Both cohorts were asked about their concerns regarding the mental health impact of the COVID-19 pandemic. Although no validated assessments of mental health were undertaken, a common theme for both cohorts was concern about increasing levels of anxiety, and the role of social distancing measures and well-being of family members in contributing to such anxiety. Similarly, a qualitative study examining the mental health impact of social distancing measures suggested that deleterious effects were evident within 5–12 days of such measures being in place.^6^

So it would seem likely that we should brace ourselves for a significant increase in mental health difficulties associated with the pandemic. But who might be at greatest risk? Individuals at increased risk of the disease and/or adverse outcomes might be expected to experience greater mental health difficulties. Several such groups have now been identified. For example, the death rate is known to be higher in men and older individuals.^7,8^The latter also being more likely to have co-existing conditions and be socially-isolated through shielding. The ethnic diversity of countries such as the US and UK has also highlighted that individuals from Black, Asian and Minority Ethnic (BAME) backgrounds appear to be affected disproportionately by the disease.^9^ Finally, recent UK data suggest that some key-workers, in particular those in social care, are at greater risk of COVID-19 related mortality.^7^ This combined with evidence from China of increased anxiety and depression among health care workers,^3^ suggests that key-workers may also be at greater risk of adverse mental health outcomes.

The aforementioned factors are, however, largely non-modifiable. Do modifiable risk factors exist which could be targets for intervention? Stress and coping theory.^10^ attests that emotional responses to challenging situations vary according to both our appraisal of stressors and the availability of psychological and social resources. Cognitions are central to our appraisal process and, in the current context, one might expect that individuals who appraise that they are at greatest risk of disease may have poorer mental health outcomes. Evidence from the Netherlands during the H1N1 pandemic supports this hypothesis, with prospective data indicating that levels of anxiety declined as perceptions of the risk of acquiring severe disease declined.^11^ Another relevant feature of our appraisal process concerns the extent to which we attend to and worry about threats to our health. This is particularly pertinent in contexts where individuals receive health threat information from the media, as in the current pandemic.^12^ Early evidence of this mechanism has emerged in the COVID-19 pandemic: with frequency of exposure to information about COVID-19 on social media found to be positively associated with mental health outcomes.^13^

In terms of resources, social support, and its corollary loneliness, are among the best established determinants of our emotional responses to stressors. Successive systematic reviews demonstrate not only poorer mental health outcomes, but also increased morbidity and mortality in individuals who perceive themselves to be more lonely and lacking in support.^14,15^A determinant also attracting attention is positive mood. It is now recognised that positive mood is not just the opposite of negative mood, but may in fact confer direct effects on well-being as well as protective effects in challenging situations.^10,16^ In terms of mental health outcomes, evidence suggests that the existence of positive mood reduces the risk of mood disorders by 28% and anxiety disorders by 53%, and also influences recovery from some mental health conditions.^17,18^

Taken together there is an urgent need to report evidence on the prevalence of mental health problems during the COVID-19 pandemic, to understand who may be at greatest risk, and to explore the psychological and social resources that may mitigate this risk. To that end, we report cross sectional findings from a community cohort survey conducted between 3^rd^ and 30^th^ April 2020 which coincided with the first 4–6 weeks of social distancing measures being introduced in the UK.

## Methods

### Recruitment and Eligibility

Ethical approval was granted from the University of Nottingham Faculty of Medicine and Health Sciences (ref: 506–2003) and the NHS Health Research Authority (ref: 20/HRA/1858). The study was launched on 3/4/20 with participants recruited in the community through a social and mainstream media campaign involving, but not limited to, Facebook and Twitter. In addition, HRA regulatory approval enabled us to approach NHS organisations and request they advertise the research through their routine communications. Recruitment continued until 30/4/20. All media directed potential participants to the study website (http://www.covidstressstudy.co.uk) through which they accessed the information sheet, consent form and online survey.

Eligibility criteria specified that participants should be: aged 18 and over; able to give informed consent; able to read English; residing in the UK at the time of completing the survey and able to provide a sample of hair at least 1 cm long. The latter was collected for the determination of the stress biomarker cortisol which will be the subject of future manuscripts.

### Procedures

Consenting participants completed an online survey implemented through JISC Online Survey (https://www.onlinesurveys.ac.uk/). This included validated measures capturing the mental health outcomes: anxiety(α=0·88), depression (α=0·76) (α#x003D;0·92) and stress (α=0·76).^19–22^ We also measured modifiable and non-modifiable variables we hypothesised would be related to these mental health outcomes due to being (i) associated with an increased risk of contracting COVID-19 and/or adverse disease outcomes; or (ii) known to be directly associated with adverse mental health outcomes. These were: age, gender, ethnicity, key-worker status, living alone, positive mood, worry about contracting COVID-19 and perceived loneliness and risk of COVID-19 (see supplementary appendix).

### Statistical analysis

We first summarised the outcome variables (depression, anxiety and stress) and participant characteristics with appropriate summary statistics and examined histograms and scatterplots. To explore the associations between non-modifiable and modifiable explanatory factors on outcome variables we conducted univariable linear regression analyses (see supplementary appendix). Multivariable linear regression analyses were then used to explore the independent relationships of non-modifiable factors (age, gender, ethnicity, keyworker status, living alone) on outcome variables. Then, in subsequent models, modifiable explanatory factors (perceived loneliness, perceived risk of COVID-19, positive mood, worry about contracting COVID-19) were added to examine the additional and independent contribution of these factors to explaining variation in the outcome variables. The variable assessing COVID-19 worry was treated as a categorical variable in all models, with “occasional worry” treated as the reference value as this was the most common response. Assumptions of linear regression (normality and homoscedasticity of residuals, linearity with continuous variables) and presence of outliers were assessed graphically. Square root transformations were used for depression and anxiety scores to satisfy assumptions. Robustness of the models were examined by removing data points with large residuals (<-3 or > 3) and comparing results to the original models. In the vast majority of models, this had no substantive effect on interpretation. Thus these results are only mentioned where interpretation may be affected. Additionally, as perceived risk of getting COVID-19 was not assessed in those who thought they had had it (n = 519) these participants are not represented in final multivariable models. As a sensitivity analysis, models were additionally re-specified excluding this explanatory variable (see supplementary appendix).

For depression and anxiety we carried out additional analyses dichotomising according to established cut-offs (scores of 10 or greater indicating moderate or severe levels). We used multiple logistic regression to estimate odds ratios with 95% confidence intervals for their associations with non-modifiable and modifiable variables.

Statistical analyses were performed using STATA (version 16).

## Results

### Cohort characteristics

The final number of participants recruited was n=3102. Of these, five were ineligible due to being less than 18 years old. Thus, yielding n = 3097 eligible participants. The largest proportion of visitors to the website came direct to the URL (62%/n=15,218), followed by 25% (n=6068) via Facebook (the remainder through other websites). The vast majority of respondents accessed the website via a mobile phone (70%/n=17045). The survey was completed in full by 100% of those who started it, consequently there were no missing data, with the exception of age, for which 2 participants entered non-numeric values.

Table 1 summarises the main characteristics of the participants and reveals that our sample was predominantly female; with a mean age of 44 years (standard deviation = 15); with participation across the UK (albeit primarily from England) and 10%/n = 296 from minority ethnic backgrounds. Fifty percent (n = 1559) described themselves as key-workers (39%/n = 1198 identifying as working in health and social care). Twenty percent (n = 649) identified themselves as having clinical risk factors which would put them at increased or greatest risk of COVID-19.

**Table 1:** Participant Demographics (n = 3097)

|  | Participants |
| --- | --- |
|  | n (%) |
| Gender |  |
| Male | 476 (15.4%) |
| Female | 2618 (84.5%) |
| Prefer not to say | 3 (0.1%) |
| **Age groups (years)** |  |
| 18-24 | 363 (11.7%) |
| 25-34 | 528 (17.1%) |
| 35-44 | 635 (20.5%) |
| 45-54 | 687 (22.2%) |
| 55-64 | 568 (18.3%) |
| 65-74 | 254 (8.2%) |
| ≥75 | 62 (2.0%) |
| **Ethnicity** |  |
| White – British, Irish, other | 2796 (90.3%) |
| Asian/Asian British – Indian, Pakistani, Bangladeshi, other | 119 (3.8%) |
| Black/Black British – Caribbean, African, other | 42 (1.4%) |
| Chinese/Chinese British | 28 (0.9%) |
| Mixed race – White and Black/Black British | 19 (0.6%) |
| Middle Eastern/Middle Eastern British – Arab, Turkish, other | 23 (0.7%) |
| Mixed race – other | 40 (1.3%) |
| Other ethnic group | 25 (0.8%) |
| Prefer not to say | 5 (0.2%) |
| **Relationship status** |  |
| Single, never married | 574 (18.5%) |
| Single, divorced or widowed | 263 (8.5%) |
| In a relationship/married but living apart | 254 (8.2%) |
| In a relationship/married and cohabiting | 1981 (64.0%) |
| Prefer not to say | 25 (0.8%) |
| **Education (highest level of attainment)** |  |
| No qualifications | 33 (1.1%) |
| Completed GSCE/CSE/O-levels or equivalent | 252 (8.1%) |
| Completed post-16 vocational course | 101 (3.3%) |
| A-levels or equivalent (at school until aged 18) | 403 (13.0%) |
| Undergraduate degree or professional qualification | 1306 (42.2%) |
| Postgraduate degree | 976 (31.5%) |
| Prefer not to say | 26 (0.8%) |
| **Place of residence** |  |
| South West England | 241 (7.8%) |
| East Midlands | 762 (24.6%) |
| Yorkshire and Humber | 293 (9.5%) |
| North East | 147 (4.8%) |
| East of England | 153 (4.9%) |
| North West | 357 (11.5%) |
| South East England | 415 (13.4%) |
| Greater London | 329 (10.6%) |
| West Midlands | 165 (5.3%) |
| Northern Ireland | 8 (0.3%) |
| Wales | 73 (2.4%) |
| Scotland | 154 (5.0%) |
| **Key-worker status** |  |
| Health, social care or relevant related support worker | 1198 (38.7%) |
| Teacher or childcare worker still travelling in to work | 70 (2.3%) |
| Transport worker still travelling in to work | 1 (0.03%) |
| Food chain worker (e.g. production, sale, delivery) | 33 (1.1%) |
| Key public services worker (e.g. justice staff, religious staff, public service journalist or mortuary worker) | 22 (0.7%) |
| Local or national government worker delivering essential public services | 41 (1.3%) |
| Utility worker (e.g. energy, sewerage, postal service) | 5 (0.2%) |
| Public safety or national security worker | 11 (0.4%) |
| Worker involved in medicines or protective equipment production or distribution | 10 (0.3%) |
| Other key worker role not listed | 168 (5.4%) |
| Not a key worker | 1538 (49.7%) |
| **Living alone (or with others)** |  |
| Living alone | 406 (13.1%) |
| Living with others | 2691 (86.9%) |
| **COVID-19 risk status** |  |
| Most at risk (e.g. suffering from advanced cancer, severe asthma/COPD, etc.) | 121 (3.9%) |
| At increased risk (e.g., being pregnant, aged over 70, etc.) | 528 (17.1%) |
| Not at-risk | 2448 (79.0%) |

### Mental health outcomes

Table 2 summarises findings in relation to levels of stress, anxiety and depression in the cohort. The mean values for all measures indicate levels that are higher in women than men and decrease with age. Overall mean values are significantly higher than previously reported population norms^23–25^. For both anxiety and depression the means for the cohort were higher for both genders compared with their respective population norms, and also for all age ranges between 25–64 years. In contrast, both men and women aged over 65 years had anxiety and depression scores consistent with previous population norms. The data suggested no significant differences in stress scores by gender, despite the combined mean score exceeding the population norm.

**Table 2:**
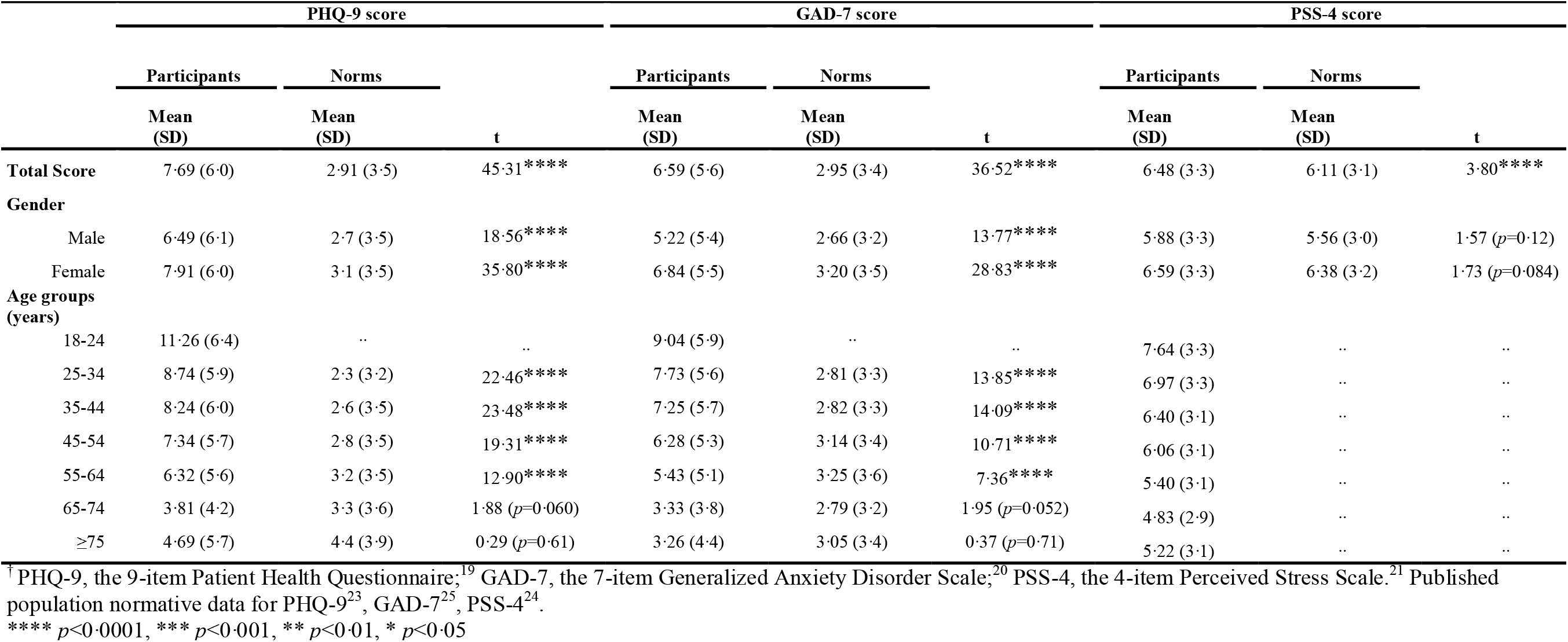
Depression (PHQ-9), anxiety (GAD-7) and stress (PSS-4) scores and published population normative data^†^.

|  | PHQ-9 score |  |  | GAD-7 score |  |  | PSS-4 score |  |  |
| --- | --- | --- | --- | --- | --- | --- | --- | --- | --- |
|  | Participants | Norms | t | Participants | Norms | t | Participants | Norms | t |
|  | Mean (SD) | Mean (SD) |  | Mean (SD) | Mean (SD) |  | Mean (SD) | Mean (SD) |  |
| **Total Score** | 7.69 (6.0) | 2.91 (3.5) | 45.31**** | 6.59 (5.6) | 2.95 (3.4) | 36.52**** | 6.48 (3.3) | 6.11 (3.1) | 3.80**** |
| **Gender** |  |  |  |  |  |  |  |  |  |
| Male | 6.49 (6.1) | 2.7 (3.5) | 18.56**** | 5.22 (5.4) | 2.66 (3.2) | 13.77**** | 5.88 (3.3) | 5.56 (3.0) | 1.57 (*p*=0.12) |
| Female | 7.91 (6.0) | 3.1 (3.5) | 35.80**** | 6.84 (5.5) | 3.20 (3.5) | 28.83**** | 6.59 (3.3) | 6.38 (3.2) | 1.73 (*p*=0.084) |
| **Age groups (years)** |  |  |  |  |  |  |  |  |  |
| 18-24 | 11.26 (6.4) | .. | .. | 9.04 (5.9) | .. | .. | 7.64 (3.3) | .. | .. |
| 25-34 | 8.74 (5.9) | 2.3 (3.2) | 22.46**** | 7.73 (5.6) | 2.81 (3.3) | 13.85**** | 6.97 (3.3) | .. | .. |
| 35-44 | 8.24 (6.0) | 2.6 (3.5) | 23.48**** | 7.25 (5.7) | 2.82 (3.3) | 14.09**** | 6.40 (3.1) | .. | .. |
| 45-54 | 7.34 (5.7) | 2.8 (3.5) | 19.31**** | 6.28 (5.3) | 3.14 (3.4) | 10.71**** | 6.06 (3.1) | .. | .. |
| 55-64 | 6.32 (5.6) | 3.2 (3.5) | 12.90**** | 5.43 (5.1) | 3.25 (3.6) | 7.36**** | 5.40 (3.1) | .. | .. |
| 65-74 | 3.81 (4.2) | 3.3 (3.6) | 1.88 (*p*=0.060) | 3.33 (3.8) | 2.79 (3.2) | 1.95 (*p*=0.052) | 4.83 (2.9) | .. | .. |
| ≥75 | 4.69 (5.7) | 4.4 (3.9) | 0.29 (*p*=0.61) | 3.26 (4.4) | 3.05 (3.4) | 0.37 (*p*=0.71) | 5.22 (3.1) | .. | .. |

<sup>†</sup> PHQ-9, the 9-item Patient Health Questionnaire;<sup>19</sup> GAD-7, the 7-item Generalized Anxiety Disorder Scale;<sup>20</sup> PSS-4, the 4-item Perceived Stress Scale.<sup>21</sup> Published population normative data for PHQ-9<sup>23</sup>, GAD-7<sup>25</sup>, PSS-4<sup>24</sup>.
\*\*\*\* *p*<0.0001, \*\*\* *p*<0.001, \*\* *p*<0.01, \* *p*<0.05

Table 3 shows the categorisation of participants in line with established cut-offs for anxiety and depression. This shows 64% of participants reported symptoms of depression and 57% reported symptoms of anxiety. When considering the thresholds at which someone would qualify for high intensity psychological support (score of 10 or greater) in the NHS,^26^ we observe that 31.6% reported moderate to severe depression and 26% moderate to severe anxiety.

**Table 3:** Prevalence of depressive and anxiety cases^†^.

|  |  | Whole sample |  | Male |  | Female |  |
| --- | --- | --- | --- | --- | --- | --- | --- |
|  | Categories | n | % | n | % | n | % |
| Depression (PHQ-9<sup>‡</sup>) | *No-Minimal Depression (0-4)* | 1125 | 36.3 | 230 | 48.3 | 894 | 34.1 |
|  | *Mild Depression (5-9)* | 994 | 32.1 | 125 | 26.3 | 868 | 33.2 |
|  | *Moderate Depression (10-14)* | 525 | 17.0 | 64 | 13.4 | 461 | 17.6 |
|  | *Moderately Severe Depression (15-19)* | 276 | 8.9 | 35 | 7.4 | 241 | 9.2 |
|  | *Severe Depression (20-27)* | 177 | 5.7 | 22 | 4.6 | 154 | 5.9 |
| Anxiety (GAD-7<sup>‡</sup>) | *No-Minimal Anxiety (0-4)* | 1344 | 43.4 | 276 | 58.0 | 1066 | 40.7 |
|  | *Mild Anxiety (5-9)* | 947 | 30.6 | 108 | 22.7 | 839 | 32.0 |
|  | *Moderate Anxiety (10-14)* | 430 | 13.9 | 44 | 9.2 | 386 | 14.7 |
|  | *Severe Anxiety (15-21)* | 376 | 12.1 | 48 | 10.1 | 327 | 12.5 |

<sup>†</sup> Cut-offs for categories in line with published guidelines for PHQ-9<sup>23</sup> and GAD-7.<sup>25</sup>
<sup>‡</sup> PHQ-9, the 9-item Patient Health Questionnaire;<sup>19</sup> GAD-7, the 7-item Generalized Anxiety Disorder Scale.<sup>20</sup>

### Individuals at greatest risk of mental health problems: associations with age, gender, ethnicity, living alone and key-worker status

When non-modifiable explanatory variables were included in a multivariable model (Table 4), we observed that for depression (square-root transformed scores), being younger (B=–0·28, 95% CI:-0·31, –0·25 per decade), female (B=0·36, 95% CI: 0·25, 0·47) and living alone (B=0·34, 95% CI: 0·25, 0·47) were all independently significantly associated with greater levels of depression. This model accounted for approximately 13% of the variance in depression scores. These results were replicated when considering depression as a binary outcome (i.e., cases requiring high intensity intervention versus not) with females having a 49% increased odds of depression and living alone associated with a 55% increase.

**Table 4:** Regression models showing associations between non-modifiable explanatory variables and depression scores.

| | B | 95% CI Lower | 95% CI Upper | $\beta$ | p |
| --- | --- | --- | --- | --- | --- |
| **PHQ-9 Total Score <sup>a</sup>** |  |  |  |  |  |
| Age (per decade) | -0.28 | -0.31 | -0.25 | -0.35 | <.0001**** |
| Female | 0.36 | 0.25 | 0.47 | 0.11 | <.0001**** |
| Live alone | 0.34 | 0.22 | 0.46 | 0.09 | <.0001**** |
| BAME background | 0.03 | -0.11 | 0.17 | 0.01 | .64 |
| Key-worker | 0.06 | -0.02 | 0.14 | 0.02 | .15 |
| **Adjusted R<sup>2</sup>=0.13, n=3090** |  |  |  |  |  |
| | Odds Ratio | 95% CI Lower | 95% CI Upper | $\beta$ | p |
| **PHQ-9 “Cases” <sup>b</sup>** |  |  |  |  |  |
| Age (per decade) | 0.67 | 0.63 | 0.71 | -1.30 | <.0001**** |
| Female | 1.49 | 1.18 | 1.89 | 0.31 | .00079*** |
| Live alone | 1.55 | 1.23 | 1.97 | 0.32 | .00025*** |
| BAME background | 1.14 | 0.88 | 1.48 | 0.08 | .32 |
| Key-worker | 1.14 | 0.97 | 1.33 | 0.14 | .11 |
| **Pseudo R<sup>2</sup>=0.06, n=3090** |  |  |  |  |  |

\*\*\*\* $p<0.0001$ , \*\*\* $p<0.001$ , \*\* $p<0.01$ , \* $p<0.05$
<sup>a</sup> A square-root transformation was applied to the dependent variable.
<sup>b</sup> a “case” is defined as a PHQ-9 score greater than or equal to 10, at which level someone would qualify for high intensity psychological support in the NHS.

For anxiety (square-root transformed scores) being younger (B = –0·24, 95% CI: –0·27, –0·22 per decade) and female (B = 0·43, 95% CI: 0·32, 0·55) were independently significantly associated with greater levels of anxiety (Table 5). This model accounted for approximately 11% of the variance and these results were replicated when considering anxiety as a binary outcome (i.e., cases requiring high intensity intervention versus not).

**Table 5:** Regression models showing associations between non-modifiable explanatory variables and anxiety scores.

| | B | 95% CI Lower | 95% CI Upper | $\beta$ | p |
| --- | --- | --- | --- | --- | --- |
| **GAD-7 Total Score<sup>a</sup>** |  |  |  |  |  |
| Age (per decade) | -0.24 | -0.27 | -0.22 | -0.30 | <.0001**** |
| Female | 0.43 | 0.32 | 0.55 | 0.13 | <.0001**** |
| Live alone | -0.03 | -0.15 | 0.09 | -0.01 | .64 |
| BAME background | 0.02 | -0.12 | 0.16 | 0.01 | .77 |
| Key-worker | 0.08 | -0.00 | 0.16 | 0.03 | .06 |
| **Adjusted R<sup>2</sup>=0.11, n=3090** |  |  |  |  |  |
| | Odds Ratio | 95% CI Lower | 95% CI Upper | $\beta$ | p |
| **GAD-7 “Cases”<sup>b</sup>** |  |  |  |  |  |
| Age (per decade) | 0.70 | 0.66 | 0.74 | -1.23 | <.0001**** |
| Female | 1.61 | 1.25 | 2.08 | 0.39 | 0.00020*** |
| Live alone | 1.02 | 0.78 | 1.32 | 0.01 | .91 |
| BAME background | 1.15 | 0.88 | 1.51 | 0.10 | .30 |
| Key-worker | 1.13 | 0.96 | 1.34 | 0.14 | .15 |
| **Pseudo R<sup>2</sup>=0.05, n=3090** |  |  |  |  |  |

\*\*\*\* $p < 0.0001$ , \*\*\* $p < 0.001$ , \*\* $p < 0.01$ , \* $p < 0.05$
<sup>a</sup> A square-root transformation was applied to the dependent variable.
<sup>b</sup> a “case” is defined as a GAD-7 score greater than or equal to 10, at which level someone would qualify for high intensity psychological support in the NHS.

For stress, being younger (B=–0·54, 95% CI: –0·61, –0·46 per decade), female (B = 0·78, 95% CI: 0·46, 1·09), living alone (B = 0·48, 95% CI: 0·15, 0·82), being from a BAME background (B = 0·45, 95% CI: 0·07, 0·84), were all independently significantly associated with greater stress; while being a key-worker was independently significantly associated with a lower stress (B = –0·24, 95% CI: –0·47, –0·02). Together the model accounted for approximately 7% of the variance in stress scores (Table 6).

**Table 6:** Regression model showing associations between non-modifiable explanatory variables and stress scores.

| | B | 95% CI Lower | 95% CI Upper | $\beta$ | p |
| --- | --- | --- | --- | --- | --- |
| **PSS-4 Total Score** |  |  |  |  |  |
| Age (per decade) | -0.54 | -0.61 | -0.46 | -0.25 | <.0001**** |
| Female | 0.78 | 0.46 | 1.09 | 0.09 | <.0001**** |
| Live alone | 0.48 | 0.15 | 0.82 | 0.05 | 0.0049** |
| BAME background | 0.45 | 0.07 | 0.84 | 0.04 | 0.022* |
| Key-worker | -0.24 | -0.47 | -0.02 | -0.04 | 0.033* |
| **Adjusted R<sup>2</sup>=0.07, n=3090** |  |  |  |  |  |

\*\*\*\* $p < 0.0001$ , \*\*\* $p < 0.001$ , \*\* $p < 0.01$ , \* $p < 0.05$

### Individuals at greatest risk of mental health problems: associations with perceived risk of COVID-19, perceived loneliness, COVID-19 worry and positive mood

Table 7 shows levels of modifiable explanatory variables (perceived risk, perceived loneliness, COVID-19 worry, and positive mood) across the whole sample, as well as by gender and age-groups.

**Table 7:** Loneliness, worry about COVID-19, perceived risk of COVID-19, and positive mood.

|  | Whole sample | Gender |  | Age groups (years) |  |  |  |  |  |  |
| --- | --- | --- | --- | --- | --- | --- | --- | --- | --- | --- |
|  |  | Male | Female | 18-24 | 25-34 | 35-44 | 45-54 | 55-64 | 65-74 | ≥75 |
| Loneliness |  |  |  |  |  |  |  |  |  |  |
| Mean (SD) | 3.86 (2.7) | 3.56 (2.7) | 3.91 (2.7) | 5.36 (2.7) | 4.36 (2.7) | 3.76 (2.7) | 3.62 (2.8) | 3.47 (2.7) | 2.70 (2.1) | 2.71 (2.4) |
| Positive mood |  |  |  |  |  |  |  |  |  |  |
| Mean (SD) | 18.99 (5.1) | 19.76 (5.1) | 18.85 (5.0) | 17.67 (4.9) | 18.82 (5.1) | 18.67 (5.0) | 18.92 (5.1) | 19.38 (5.0) | 20.72 (4.6) | 21.56 (5.2) |
| Perceived risk of COVID-19 |  |  |  |  |  |  |  |  |  |  |
| Mean (SD) | 4.75 (2.2) | 4.46 (2.2) | 4.80 (2.2) | 4.10 (2.0) | 4.92 (2.2) | 5.15 (2.2) | 5 (2.2) | 4.78 (2.3) | 4.21 (2.1) | 3.31 (1.9) |
| Worry about COVID-19 |  |  |  |  |  |  |  |  |  |  |
| No worry (n, %) | 512 (16.5%) | 105 (22.1%) | 406 (15.5%) | 105 (28.9%) | 108 (20.5%) | 91 (14.3%) | 91 (13.3%) | 65 (11.4%) | 39 (15.4%) | 13 (21.0%) |
| Occasional worry (n, %) | 2050 (66.2%) | 318 (66.8%) | 1731 (66.1%) | 208 (57.3%) | 320 (60.6%) | 427 (67.2%) | 466 (67.8%) | 396 (69.7%) | 188 (74.0%) | 45 (72.6%) |
| Much worry (n, %) | 413 (13.3%) | 40 (8.4%) | 373 (14.3%) | 39 (10.7%) | 77 (14.6%) | 91 (14.3%) | 94 (13.7%) | 85 (15.0%) | 24 (9.5%) | 3 (4.8%) |
| Most worry (n, %) | 122 (3.9%) | 13 (2.7%) | 108 (4.1%) | 11 (3.0%) | 23 (4.4%) | 26 (4.1%) | 36 (5.2%) | 22 (3.9%) | 3 (1.2%) | 1 (1.6%) |

When modifiable explanatory variables were added into the multivariable model for depression: this revealed that greater perceived loneliness (B = 0·10, 95% CI: 0·09, 0·12), lower positive mood (B = –0·12, 95% CI: –0·12, –0·11) and greater than occasional worry about getting COVID-19 (much of time: B = 0·28, 95% CI: 0·18, 0·38; most of time: B = 0·32, 95% CI: 0·13, 0·50), were all independently and significantly associated with greater levels of depression, in addition to age and gender. The model accounted for approximately 56% of the variance in depression scores. While perceived risk of COVID-19 was not statistically significant, in sensitivity analyses where large residuals were excluded (<-3/>3) this became statistically significant (B = 0·02, 95% CI: 0·00, 0·03). These results were largely replicated when considering depression as a binary outcome although gender was no longer statistically significant (Table 8).

**Table 8:**
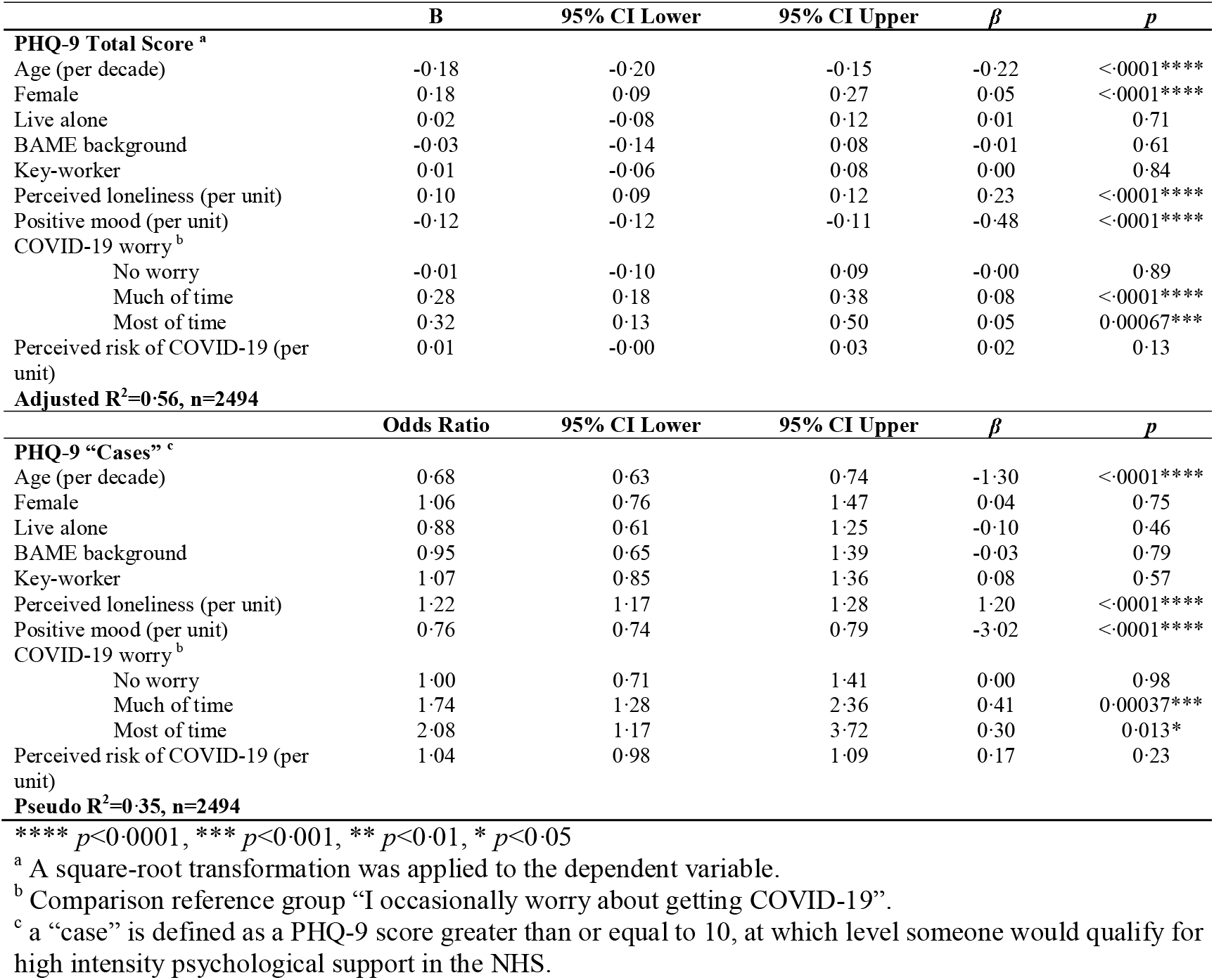
Regression models showing associations between modifiable explanatory variables and depression scores.

For anxiety, the model revealed that greater perceived loneliness (B = 0·06, 95% CI: 0·04, 0·07), lower positive mood (B = –0·12, 95% CI: –0·13, –0·11) and greater perceived risk of COVID-19 (B = 0·04, 95% CI: 0·02, 0·05) were all independently and significantly associated with greater anxiety, in addition to the non-modifiable factors of being younger, female and living alone. Further, those participants who experienced greater than occasional worry about getting COVID-19 were significantly more likely to have higher levels of anxiety (much of time: B = 0·58, 95% CI: 0·47, 0·68; most of time: B = 0·87, 95% CI: 0·68, 1·06); with those who did not worry at all about getting COVID-19 being likely to have lower anxiety (B = –0·19, 95% CI: –0·28, –0·09). The model accounted for approximately 54% of the variance in anxiety scores. These results were largely replicated when considering anxiety as a binary outcome, although gender and not worrying at all about getting COVID-19 were no longer statistically significant (Table 9).

**Table 9:** Regression models showing associations between modifiable explanatory variables and anxiety.

|  | **B** | **95% CI Lower** | **95% CI Upper** | **$\beta$** | ***p*** |
| --- | --- | --- | --- | --- | --- |
| **GAD-7 Total Score <sup>a</sup>** |  |  |  |  |  |
| Age (per decade) | -0.16 | -0.18 | -0.13 | -0.19 | <.0001**** |
| Female | 0.25 | 0.15 | 0.34 | 0.07 | <.0001**** |
| Live alone | -0.25 | -0.35 | -0.15 | -0.07 | <.0001**** |
| BAME background | -0.08 | -0.20 | 0.03 | -0.02 | 0.17 |
| Key-worker | -0.04 | -0.11 | 0.03 | -0.02 | 0.27 |
| Perceived loneliness (per unit) | 0.06 | 0.04 | 0.07 | 0.12 | <.0001**** |
| Positive mood (per unit) | -0.12 | -0.13 | -0.11 | -0.48 | <.0001**** |
| COVID-19 worry <sup>b</sup> |  |  |  |  |  |
| No worry | -0.19 | -0.28 | -0.09 | -0.05 | 0.00015*** |
| Much of time | 0.58 | 0.47 | 0.68 | 0.16 | <.0001**** |
| Most of time | 0.87 | 0.68 | 1.06 | 0.13 | <.0001**** |
| Perceived risk of COVID-19 (per unit) | 0.04 | 0.02 | 0.05 | 0.06 | <.0001**** |
| **Adjusted R<sup>2</sup>=0.54, n=2494** |  |  |  |  |  |
|  | **Odds Ratio** | **95% CI Lower** | **95% CI Upper** | **$\beta$** | ***p*** |
| **GAD-7 “Cases” <sup>c</sup>** |  |  |  |  |  |
| Age (per decade) | 0.69 | 0.63 | 0.75 | -1.34 | <.0001**** |
| Female | 1.17 | 0.82 | 1.67 | 0.14 | 0.37 |
| Live alone | 0.67 | 0.46 | 0.99 | -0.32 | 0.044* |
| BAME background | 0.96 | 0.65 | 1.43 | -0.03 | 0.85 |
| Key-worker | 0.90 | 0.70 | 1.15 | -0.13 | 0.40 |
| Perceived loneliness (per unit) | 1.11 | 1.06 | 1.17 | 0.67 | <.0001**** |
| Positive mood (per unit) | 0.77 | 0.75 | 0.80 | -3.07 | <.0001**** |
| COVID-19 worry <sup>b</sup> |  |  |  |  |  |
| No worry | 0.75 | 0.52 | 1.09 | -0.24 | 0.14 |
| Much of time | 3.86 | 2.86 | 5.22 | 1.06 | <.0001**** |
| Most of time | 11.57 | 5.88 | 22.77 | 1.06 | <.0001**** |
| Perceived risk of COVID-19 (per unit) | 1.07 | 1.01 | 1.14 | 0.35 | 0.024* |
| **Pseudo R<sup>2</sup>=0.36, n=2494** |  |  |  |  |  |

\*\*\*\* $p < 0.0001$ , \*\*\* $p < 0.001$ , \*\* $p < 0.01$ , \* $p < 0.05$
<sup>a</sup> A square-root transformation was applied to the dependent variable.
<sup>b</sup> Comparison reference group “I occasionally worry about getting COVID-19”.
<sup>c</sup> a “case” is defined as a GAD-7 score greater than or equal to 10, at which level someone would qualify for high intensity psychological support in the NHS.

The multivariable model for stress scores showed that greater perceived loneliness (B=0·19, 95% CI: 0·15, 0·23), lower positive mood (B=–0·38, 95% CI:-0·40, –0·36), greater than occasional worry about getting COVID-19 (much of time: B=0·37, 95% CI: 0·10, 0·63; most of time: B=1·02, 95% CI: 0·54, 1·50), and greater perceived risk of getting COVID-19 (B=0·06, 95% CI:0·02, 0·11) were all independently and significantly associated with greater stress, in addition to being younger, female, living alone and not being a key-worker. In robustness analyses, when removing large residuals (<-3 or > 3) having a BAME background was also a statistically significant independent predictor (B=0·29, 95% CI: 0·00, 0·58). This model accounted for approximately 57% of the variance in stress scores (Table 10).

**Table 10:**
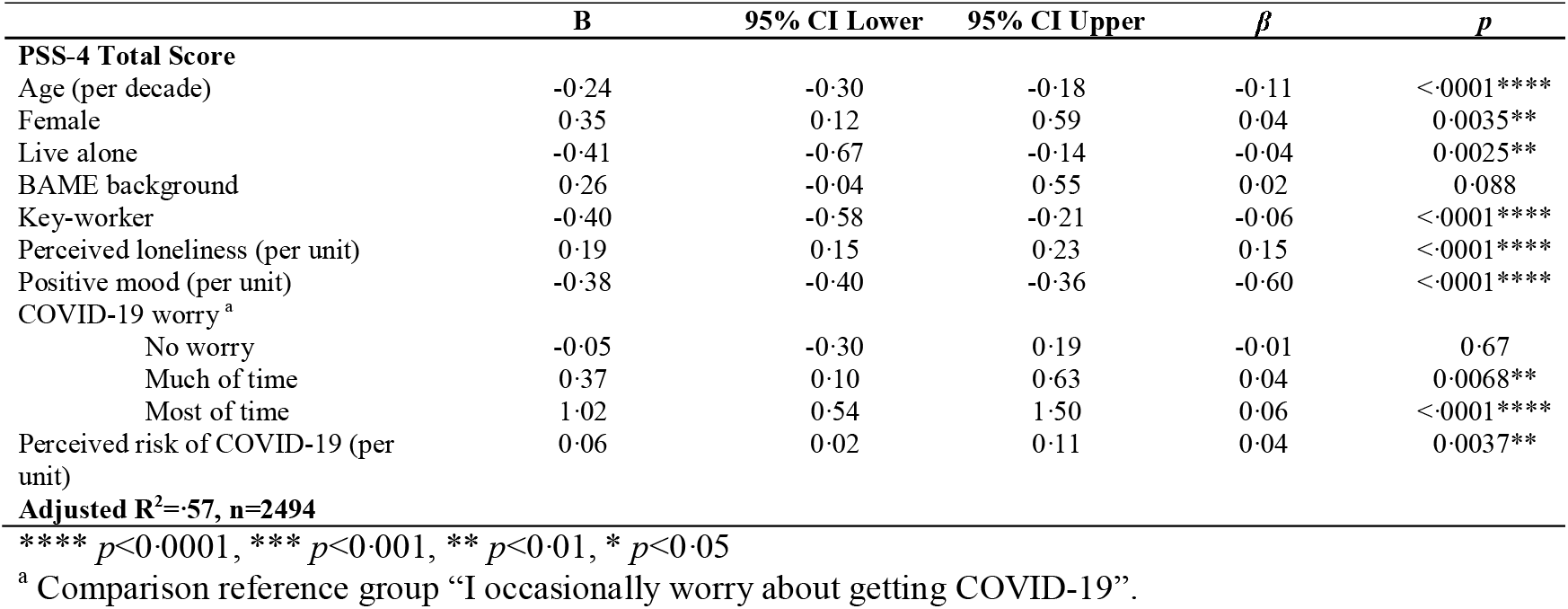
Regression model showing associations between modifiable explanatory variables and stress scores.

## Discussion

We report findings from a community cohort study established in the UK to examine the mental health consequences of the COVID-19 pandemic. Our results pertain to the experiences of people within the first four to six weeks of social distancing measures being introduced, and focus on self-reported depression, anxiety and stress scores. The findings indicated that mean levels of depression, anxiety and stress significantly exceeded recent population norms.^23–25^ Models examining the relationship between these mental health outcomes and non-modifiable explanatory factors accounted for only a modest proportion of the variance (7–13%). Increased depression was associated with being younger, female and living alone; increased anxiety was associated with being younger and female; and increased stress was associated with being younger, female, living alone, being from a BAME background and not being a keyworker. In contrast, when we added the hypothesised modifiable variables into our multivariable models we observed that they accounted for a much larger proportion of the variance (54–57%) with significant independent effects emerging for positive mood, perceived loneliness and worry about getting COVID-19 for all three outcomes, as well as perceived risk of COVID-19 emerging as significant for anxiety and stress.

These findings highlight a number of issues worthy of discussion. First, both mean scores and measures of caseness indicate that the COVID-19 pandemic is having widespread and deleterious effects on the emotional well-being of people in the UK. This is true for depression, generalised anxiety disorder and stress and is in keeping with observations from other countries.^2,3^ Indeed, the proportion of participants who would require intensive support for depression and anxiety in the NHS does not compare favourably with recent historical estimates of the prevalence of mental health problems in the UK. For example, the 2014 ONS report on adult psychiatric morbidity reported a prevalence of 17% for six different common mental disorders.^27^ The prevalence of depression alone in the context of this pandemic is almost double this.

Second, the non-modifiable explanatory variables significantly associated with all three of our mental health outcomes were being younger and being female. These findings are consistent with unpublished data from another UK community cohort recruited during the COVID-19 pandemic with a similar gender profile to our own,^28^ suggesting that these groups may be the most in need of intervention. Although this runs counter to our hypothesis that the greatest psychological morbidity would be observed in individuals at greatest risk of COVID-19, it is consistent with previous work which has shown that individual’s perceptions of disease risk are often poorly related to actual risk.^29^ Alternatively, the results may reflect the fact that the pandemic has resulted in a panoply of challenges to mental health that go beyond the disease itself. It could be hypothesised, for example, that some of the more immediate consequences such as unemployment, financial concerns and increased domestic violence would disproportionately affect younger people and women and this may explain our findings.

A third, and related issue, is that although being younger and female were consistently associated with poorer mental health, the relationship was modest, accounting for, at best, 13% of the variance. In contrast, the modifiable explanatory measures when added to the multivariable models accounted for 52–57% of the variance. These findings are encouraging as they suggest that there is considerable potential for us to develop interventions to mitigate the mental health effects of the pandemic. The measures of perceived loneliness, positive mood and worry about getting COVID-19 were strongly associated with all three outcomes and thus would be appropriate cognitions to be targeted in future interventions.^30^

A further issue concerns the effects of the pandemic beyond mental health. It is well known that when negative mood states persist over time they result in the dysregulation of physiological systems involved in the regulation of the immune system.^31^ Thus, there exists significant potential for the psychological harm inflicted by the pandemic to translate into physical harm. This could include an increased susceptibility to the virus, worse outcomes if infected, or indeed poorer responses to vaccinations in the future.^32^ Studies providing longitudinal data on the prevalence of psychological morbidity and appropriate biomarkers (e.g., cortisol) will be required to determine whether the risks to physical health go beyond the hypothetical.

Finally, we would like to acknowledge several limitations. These include the cross-sectional design of the work which impedes an analysis of cause and effect; the limited generalisability of our cohort inflicted by the self-selected community cohort design and the absence of information on pre-existing mental health conditions which are likely to impact on the severity and prevalence of psychological morbidity.^1^ Nonetheless, we are among the first to provide evidence from a large cohort on the mental health impact of the COVID-19 pandemic on people in the UK; to identify groups who may be at particular risk, as well as potential targets for therapeutic intervention.

## Data Availability

Data will be deposited in the University of Nottingham data archive. Access to this dataset will be embargoed for a period of 12 months in order for us to complete further analysis of the dataset. After that it may be shared with the consent of the Chief Investigator.

## Acknowledgements

KA was supported by funding from the National Institute for Health Research School for Primary Care Research (NIHR SPCR). The views expressed are those of the author(s) and not necessarily those of the NIHR, the NHS or the Department of Health.

TC acknowledges the financial support of the Department of Health via the National Institute for Health Research (NIHR) Specialist Biomedical Research Centre for Mental Health award to the South London and Maudsley NHS Foundation Trust (SLaM) and the Institute of Psychiatry at King’s College London. The views expressed are those of the authors and not necessarily those of the NHS, the NIHR or the Department of Health and Social Care.

## Author contributions

Ru Jia: study design, coordination and management of recruitment, preparation, analysis and interpretation of data and preparation of manuscript.

Kieran Ayling: study design, coordination and management of recruitment, preparation, analysis and interpretation of data and preparation of manuscript.

Trudie Chalder: study design, analysis and interpretation of data and preparation of manuscript

Adam Massey: study design, coordination and management of recruitment, preparation, analysis and interpretation of data and preparation of manuscript.

Elizabeth Broadbent: study design, interpretation of data and preparation of manuscript

Carol Coupland: study design, analysis and interpretation of data and preparation of manuscript

Kavita Vedhara: research lead, study design, coordination and management of recruitment, preparation, analysis and interpretation of data and preparation of manuscript.

As corresponding author, KV had access to all the data in the study and had final responsibility for the decision to submit for publication

